# Mucin-7 as a Potential Candidate Risk Allele for Cleft Lip and/or Palate

**DOI:** 10.1101/2024.04.15.24305736

**Authors:** Rachel A. Montel, Tatiana P. Rengifo, Sulie L. Chang

## Abstract

Cleft lip and/or palate (CL/P) occur in approximately 1 in 700 live births in the United States. High hereditary rates (50-80%) of CL/P indicate a strong genetic cause. The concept of strong genetic causes has been well-demonstrated in previous studies such as GWAS studies that identified *IRF6* for Van der Woode syndrome. Since the risk for genetic factors is strongly associated with CL/P, we hypothesized that RNA sequencing (RNA-seq) from CL/P patients may reveal enriched genes. Differential expression analysis examined changes in gene expression in CL/P patients compared to healthy controls, and gene-enrichment in relevant pathways. To explore the relationship between variants driving the observed changes in gene expression, we performed variant analysis and reported all CL/P-specific single nucleotide polymorphisms (SNPs). Our findings demonstrate that the normally upregulated *MUC7* gene is significantly downregulated in CL/P patients. Using our list of prioritized differentially expressed genes (DEGs), we observed significantly enriched pathways for biological processes related to cornification, skin and epidermis development, and keratinocyte and epidermal cell differentiation. By performing variant analyses, a single nucleotide polymorphism (SNP) in *MUC7*, and 47 SNPs in 20 additional genes (*CLCA4*, *ETNK2*, *ERLNC1*, *HAL*, *HOPX*, *IVL*, *KLK11*, *LIPK*, *LY6D*, *MUC21*, *NCCRP1*, *NEBL*, *PHYH*, *SERPINB11*, *SERPINB4*, *SORD*, *SPINK5*, *SULT2B1*, *TMEM154*, *TMPRSS11A*) were revealed. To our knowledge, this is the first report on the potential role of *MUC7* in contributing to CL/P. Together, these findings provide further insight into the genetic causes of CL/P.

## Introduction

Clefts of the lip and/or palate (CL/P), congenital orofacial malformations, are common birth defects of the lip and/or palate, affecting 1 in 700 live births worldwide and 16.86 per 10,000 live births in the United States^1,2^. CL/P is divided into three categories: cleft lip with palate (CLP), isolated cleft lip (CL), and isolated cleft palate (CP) with CLP occurring at the highest prevalence with the worst outcomes^3^. The consequence of this complex anomaly has lifelong implications for affected individuals and requires multidisciplinary treatment^4–6^.

The etiology of CL/P is multifactorial. Genetic and environmental factors play a role with an estimated heritability rate between 50 and 80%^7–9^. CL/P research in the past few years has been marked by significant breakthroughs with studies characterizing the underlying gene defects associated with several important clefting syndromes^10–15^. Interferon regulatory factor-6 (*IRF6*) gene was shown to cause van der Woude syndrome and the poliovirus receptor related-1 (*PVRL1*) gene is responsible for an autosomal recessive ectodermal dysplasia syndrome associated with clefting^16–20^. With the use of newer technologies, genome-wide association studies (GWAS) in CL/P explained about 60 common risk loci^21^. Still, the genetic basis of CL/P remains unclear and demonstrates a significant gap in knowledge.

Given this information, we hypothesized that using the RNA sequencing (RNA-seq) data from CL/P patients could reveal the role of high impact genes that could increase the risk of CL/P^23–24^. To assess this hypothesis, we examined (1) gene expression in CL/P patients compared to healthy controls, (2) gene-enrichment in relevant pathways, and (3) single nucleotide polymorphisms (SNPs) potentially driving upregulation or downregulation in expression. Our study comprehensively examined compensations in gene expression in CL/P-related pathways. The purpose of this paper is the opportunity to use bioinformatics to explore differential changes in gene expression and SNPs contributing to the etiology of CL/P.

## Methods

### Information on sequencing data

All RNA-seq data used in this study was accessed from NCBI (SRR13662566). SRA were selected for samples related to CL/P and six patient sample SRAs were downloaded as uploaded by Zhang et al^22^. This set contains whole blood samples from 4 CL/P patients and 2 healthy controls.

### Differential expression analysis

Differential expression analyses were performed using the Advanced RNA-seq workflow in CLC Genomics Workbench^25^ and the human hG38 reference genome as map. The RNA-seq workflow was used to align the sequences of each sample against the predicted gene and transcript tracks, using the human hG38 reference genome as map. Transcript levels were quantified in reads per kilobase per mapped reads (RPKM). False discovery rate (FDR) p-values and absolute fold change for the differentially expressed genes (DEGs) were calculated. The p-values were log-transformed. Significant DEGs were visualized using the VolcaNoseR volcano plot^26^.

### Functional enrichment analysis

Canonical pathway enrichment was performed using Ingenuity Pathway Analysis (IPA) suite (version 1.0, Qiagen, German)^27^. Enrichment pathways of the DEGs were generated based on the IPA database. Significantly enriched DEGs were identified as members of pathways and functional categories using Fisher’s exact test. All relevant gene regulatory networks were observed in Ingenuity Knowledge Base.

Gene ontology was analyzed using PANTHER Classification System^28^. Several pathways were selected for their role in biological processes that are or may be related to CL/P. Ancestor charts were generated with AmiGO2^29,30^. Gene annotation was based on 1000 Genomes^31^ [internationalgenome.org], GTEx portal^32^ [www.gtexportal.org], GenAtlas^33^ [bio.tools/genatlas], and GnomAD^34^ [http://gnomad.broadinstitute.org].

### SNP variant detection

Single-variant analysis was performed using the Basic Variant Detection workflow in CLC Genomics Workbench. Enriched variants common in CL/P compared to healthy controls were identified using the Fisher exact test. All variants in protein-coding regions in CL/P samples were selected; 5’UTR and intronic variants were excluded. Variants were sorted by gene and prediction scores (Polyphen and SIFT)^35^ were reported.

## Results

In the context that CL/P has evidence of genetic susceptibility, we hypothesized gene expression may be upregulated or downregulated in CL/P individuals compared to healthy controls. Using RNA sequencing, significant changes in gene expression were observed. To further test the status of differentiation-related genes and the quantitation of gene expression, statistical analyses were performed. Compared to healthy controls, CL/P individuals showed significant upregulation for *MAL2* (p<10^-35^) and downregulation for *MUC7* (p<10^-113^) as assessed by differential expression analysis (Figure 1).

**Figure 1.**
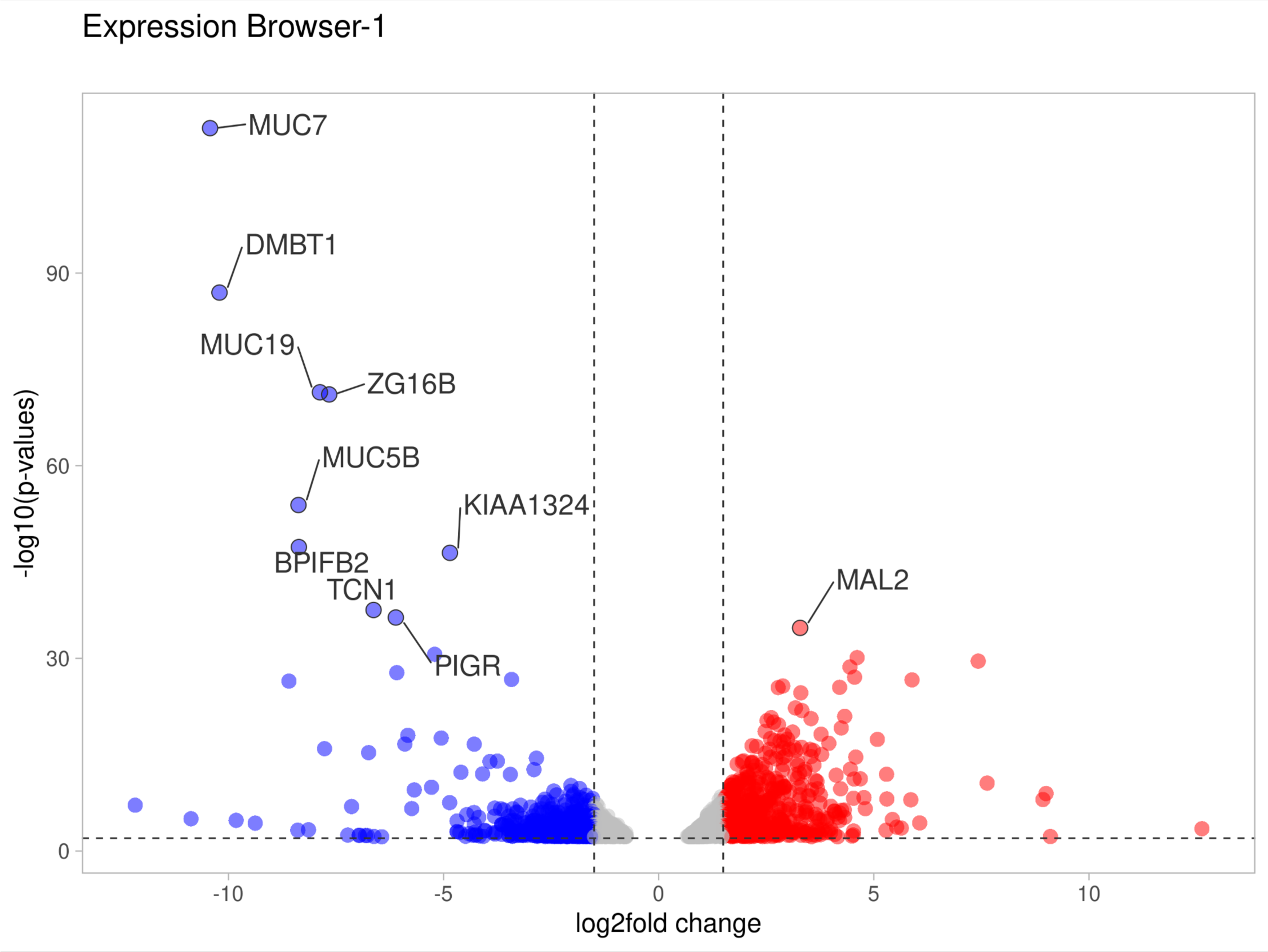
*MUC7* is differentially expressed in lip tissues of CL/P patients than healthy controls. Volcano plot showing DEGs derived from volcanoplot analysis of CL/P patients from healthy controls. Differentially expressed genes are noted (FDR<10^-28^); all upregulated (red) and downregulated (blue) genes are shown. The FDR cut-off was p-value<0.05. P-value was calculated by Fisher’s exact test.

We prioritized a set of genes (FDR p-value<10^-28^) out of 2961 differentially expressed genes (FDR p-value<0.05). Among our list of prioritized genes, the genes known to play a significant role in the development of the palate are *MUC7*, *DMBT1*, *MUC19*, *ZG16B*, *MUC5B*, *BPIFB2*, *TCN1*, *PIGR*, *MAL2*, and *SCGB3A1*. The genes contributing to development, cell regulation, or cell differentiation are *DMBT1* and *SCGB3A1*. In the family of mucin genes normally upregulated in the minor salivary glands, *MUC7*, *MUC19*, and *MUC5B* are enriched. Since the mucin genes are found in the minor salivary glands, this gene family is of high importance for CL/P. We observed two normally upregulated genes *MUC7* (p<10^-113^) and *MUC19* (p<10^-72^) were significant downregulated (Figure 1). Consistent with our interest in this gene family, we analyzed the *MUC7* pathway using IPA for overlapping between *MUC7* and other relevant genes (Figure 2). One interaction important to our findings was observed for *MUC7* and *SERPINB4*.

**Figure 2.**
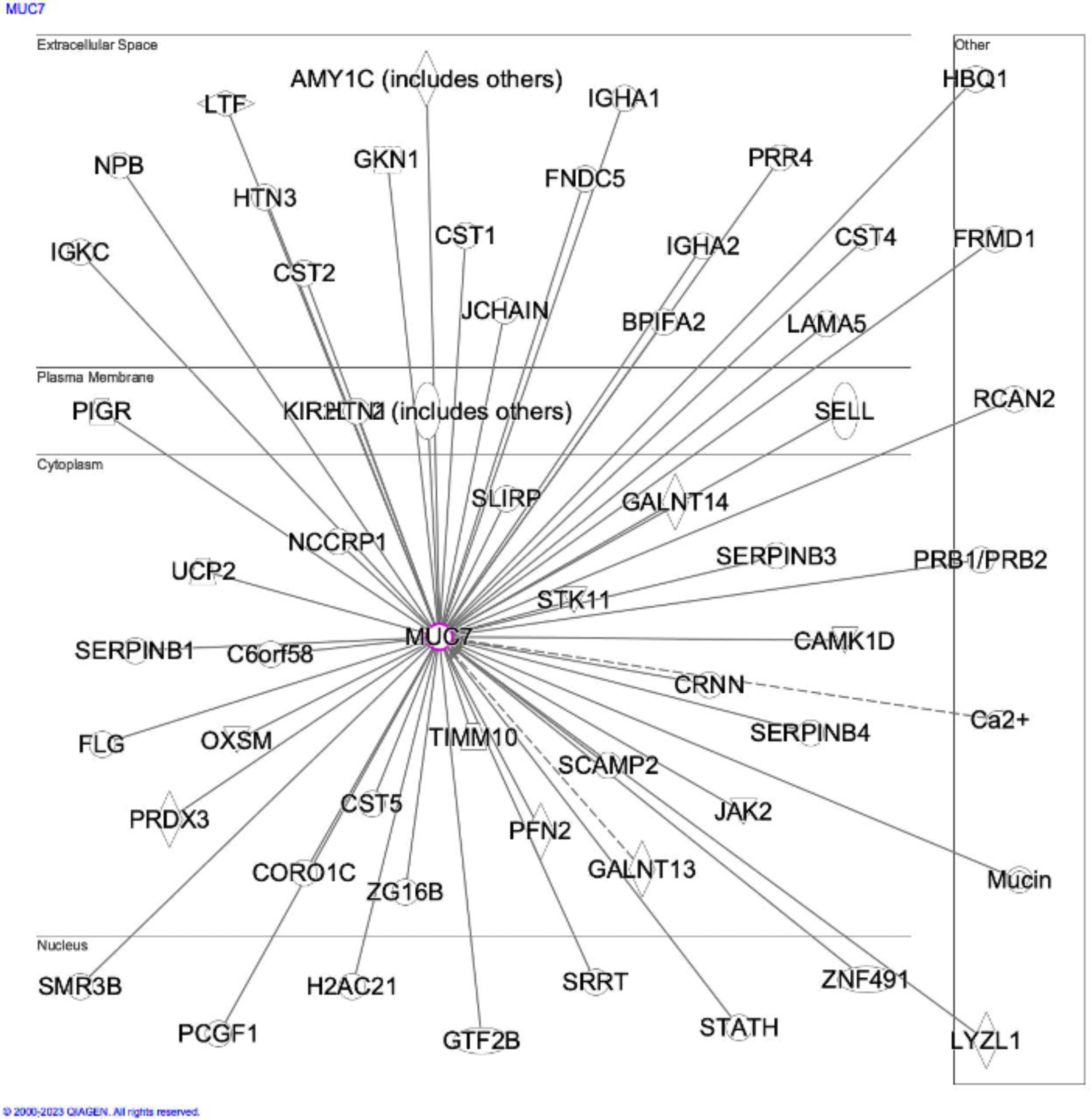
The role of *MUC7* with interacting genes. The MUC7 pathway was explored in IPA pathway analysis^27^ and organized by cellular location. The interaction of MUC7 with SERPINB4 is of particular importance since both genes contain SNPs based on our variant analysis.

Based on the concept that cleft lip and palate occurs in response to disruptions during embryogenesis and/or development, we investigated the top disease and biological functions. For the category of diseases and disorders, cancer, organismal injury and abnormalities, endocrine system disorders, gastrointestinal disease, and dermatological diseases and conditions were significantly enriched (Table 1). For the category of physiological system development and function, embryonic development, hair and skin development and function, organ development, organismal development, and tissue development were significantly enriched (Table 2). In looking at biological processes related to our differential expression set, cornification, skin development, keratinocyte differentiation, epidermis development, and epidermal cell differentiation were significantly enriched (Figure 3a). When we expanded our analysis to include molecular function, proton-transporting ATP synthase activity was significantly enriched along with other transporter activities (Figure 3b). Since cornification embodies pathways related to development and differentiation of the skin, keratin and epidermis, the cornification pathway was further explored (Figure 4). Our observations of the pathway for cornification demonstrate an important role in processes such as keratinization, epidermal cell differentiation, skin development and tissue development, that are strongly related to CL/P.

**Figure 3.**
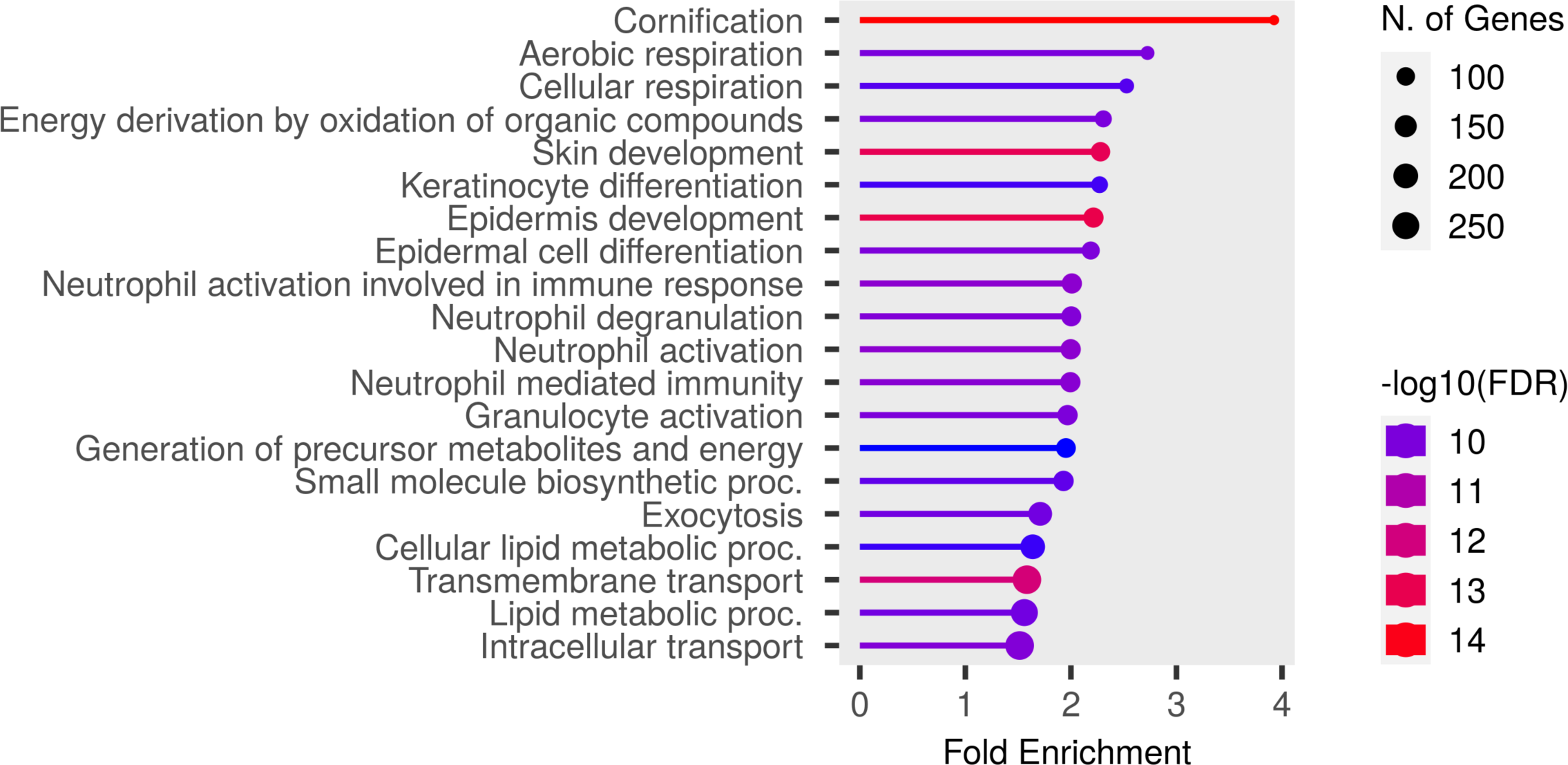
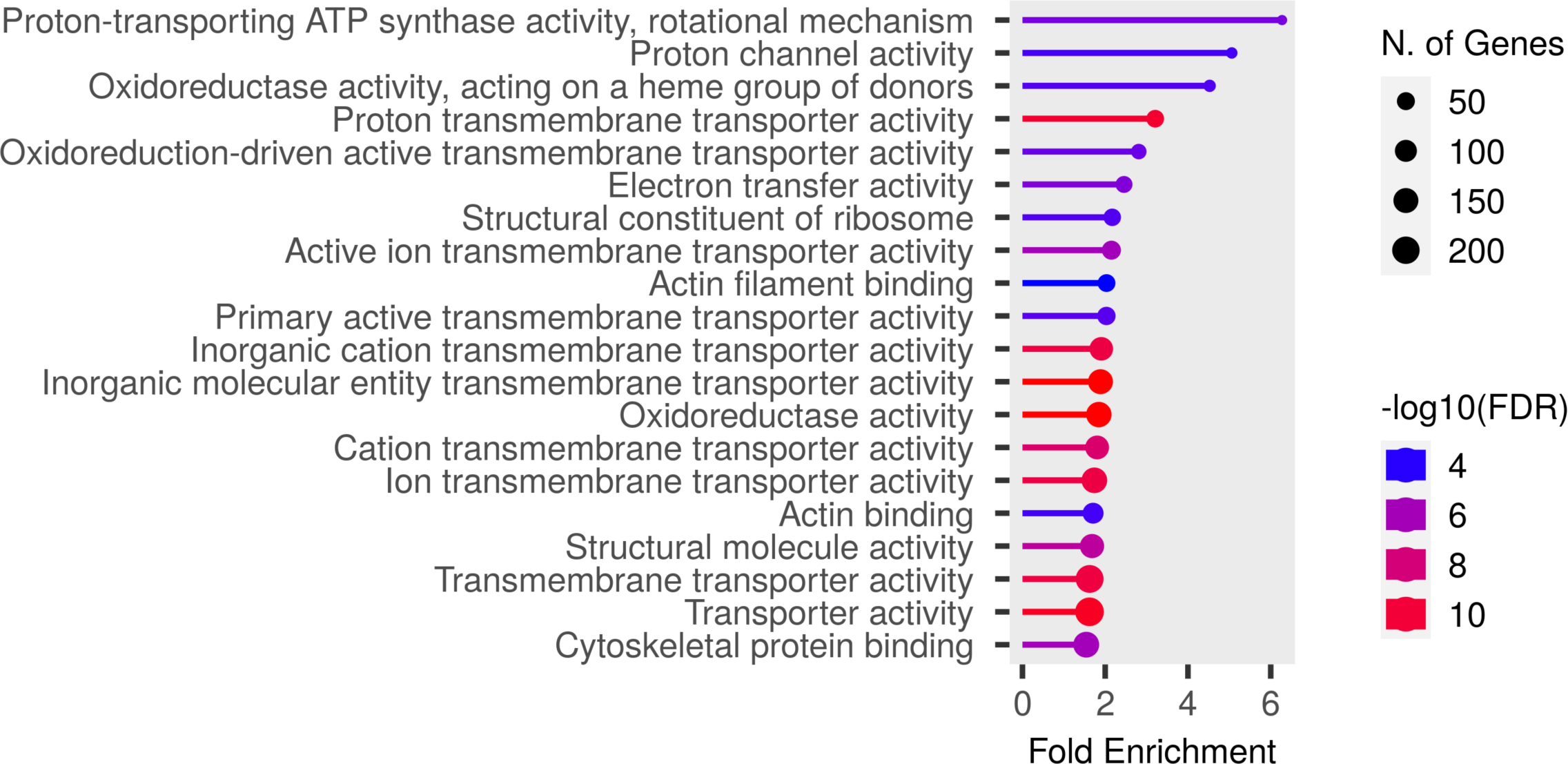
Biological process and molecular function for all differentially expressed genes. (A) Biological process for all differentially expressed genes. (B) Molecular function for all^29^ differentially expressed genes. Enrichment plot produced by ShinyGO (version 0.76.3).

**Figure 4.**
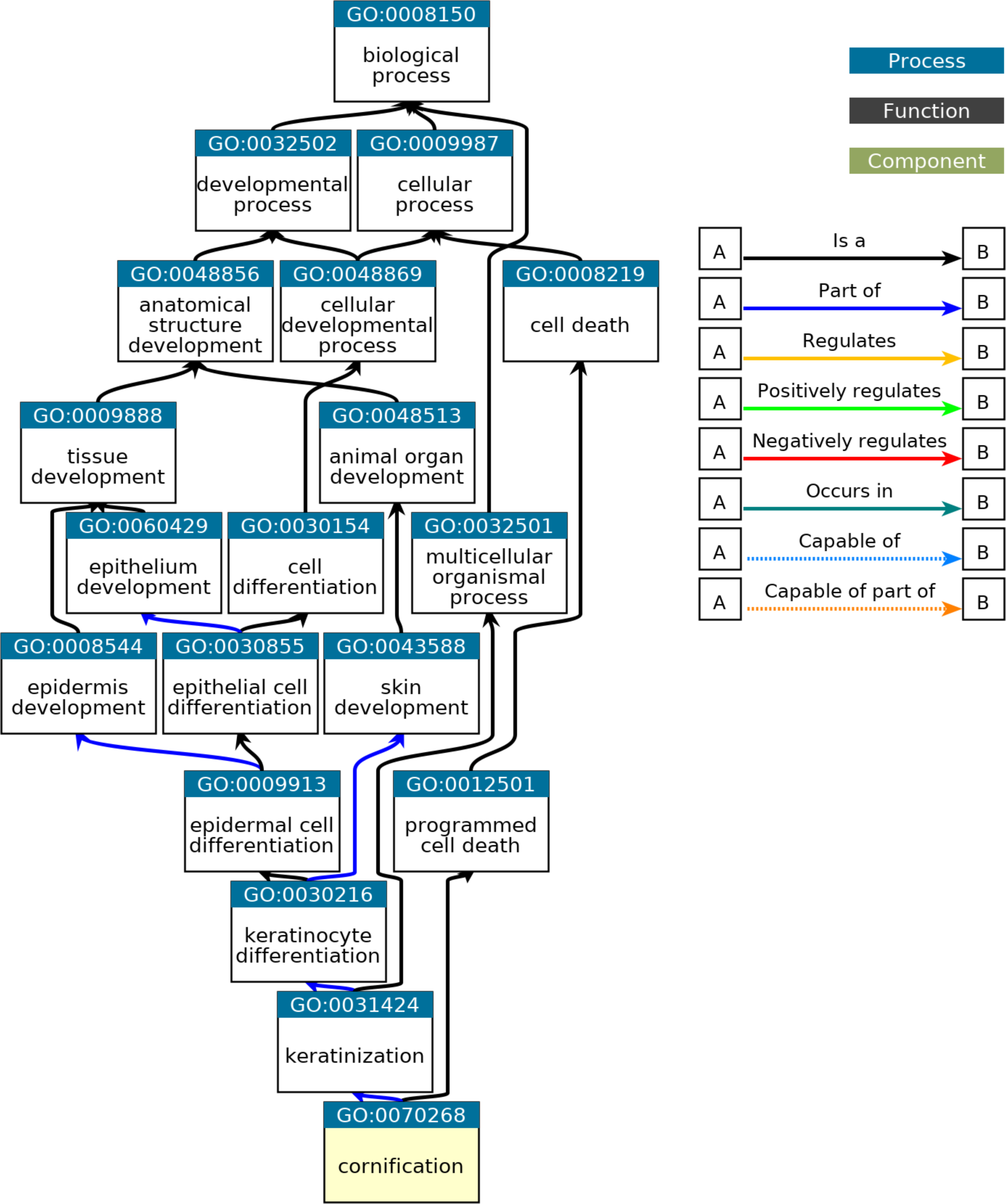
Cornification ancestor chart for significantly enriched biological processes. Gene ontology (GO) process for cornification (fold enrichment 4) demonstrating biological and cellular processes involved^30^. The processes involved in cornification include keratinization, epidermal cell differentiation, skin development and tissue development, which are strongly related to CL/P.

**Table 1.** Significantly Enriched Pathways for Physiological Systems and Functions^24^.

| **Name** | **p-value range** | **# Molecules** |
| --- | --- | --- |
| Embryonic Development | 3.98E-04 - 1.71E-11 | 187 |
| Hair and Skin Development and Function | 3.74E-04 - 1.71E-11 | 122 |
| Organ Development | 3.98E-04 - 1.71E-11 | 244 |
| Organismal Development | 4.80E-04 - 1.71E-11 | 430 |
| Tissue Development | 4.70E-04 - 1.71E11 | 287 |

**Table 2.** Significantly Enriched Pathways for Diseases and Disorders^24^.

| **Name** | **p-value range** | **# Molecules** |
| --- | --- | --- |
| Cancer | 4.71E-04 - 2.46E-139 | 2154 |
| Organismal Injury and Abnormalities | 5.02E-04 - 2.46E-139 | 2183 |
| Endocrine System Disorders | 1.22E-04 - 2.54E-39 | 1667 |
| Gastrointestinal Disease | 4.50E-04 - 3.22E-37 | 1823 |
| Dermatological Diseases and Conditions | 3.85E-04 - 6.58E-33 | 1467 |

Since changes in gene expression may be from gene variations, we investigated if any single nucleotide polymorphisms (SNPs) were associated with the status of differentiation-related genes. To assess SNPs present in CL/P individuals but absent in healthy controls, variant analysis using CLC Workbench was performed. We selected all SNPs in protein-coding regions observed only in CL/P samples and prediction scores (Polyphen, SIFT)^35^ were reported. Forty-eight SNPs were observed across 21 genes (*CLCA4*, *ETNK2*, *ERLNC1*, *HAL*, *HOPX*, *IVL*, *KLK11*, *LIPK*, *LY6D*, *MUC21*, *MUC7*, *NCCRP1*, *NEBL*, *PHYH*, *SERPINB11*, *SORD*, *SPINK5*, *SULT2B1*, *TMEM154*, *TMPRSS11A*) (Supplemental Table 1). Of the observed SNPs, 4 were predicted as possibly damaging, 6 were predicted as probably damaging, and 15 were predicted as unknown. Out of the 21 genes, SNPs were observed in *MUC7* (rs145745951) and *SERPINB4* (rs576273012); these genes were shown to interact in IPA’s *MUC7* pathway analysis (Figure 2).

## Discussion

Recent studies demonstrate that syndromes of CL/P have strong genetic links and CL/P hereditability is between 50% and 80%^7–9^. Based on these observations, we hypothesized that cleft lip and palate may have altered transcriptomic expression in RNA sequencing from CL/P patients. Our findings demonstrate that the normally upregulated *MUC7* found in the minor salivary gland is downregulated in CL/P patients. *MUC7* encodes mucin-7, a small salivary mucin found the minor salivary glands. To our knowledge, this is the first report on the association of *MUC7* with CL/P. We also observe another normally upregulated family of the mucin genes, *MUC19*, is similarly downregulated.

It is well understood that CL/P forms from disruptions during embryonic developmental processes including those of respiration, facial and dental development, and many of the syndromic genes associated with CL/P are expressed in a variety of tissues and organs^1–3^. As part of this study to assess gene-set enrichment, our analysis identified 1) significantly enriched genes whose gene expression may contribute to the risk of CL/P and 2) significant enrichment in pathways for physiological system development and function, and diseases and disorders involved in CL/P. We observed pathways related to embryonic development, hair and skin development and function, organ development, organismal development, and tissue development were enriched. These enriched pathways support the concept that explanations for risk for CL/P could lie in genes related to these pathways. Since the enriched pathways provided evidence for CL/P, we explored biological process enrichment and observed cornification was significantly enriched (fold enrichment 4). Cornification is the process in which epidermal keratinocytes undergo terminal differentiation and programmed cell death; that results in the formation of the outermost skin barrier.

Based on the concept that changes in gene expression often results from variations in SNPs, we hypothesized that SNPs would be found in the most significantly upregulated or downregulated genes. Our findings demonstrate that 10 out of 48 SNPs were potentially damaging. Based on our observations that *MUC7* is differentially expressed, a single SNP (rs145745951) was reported. The prediction of this SNP using Polyphen was ‘unknown’ indicating that further studies should be performed to characterize the influence of this SNP on mucin-7 functionality. We also observed a SNP (rs576273012) in *SERPINB4* found in the *MUC7* pathway generated in IPA.

Together, our study suggests that *MUC7* may contribute to clefting in CL/P individuals. The evidence that *MUC7* is differentially downregulated with a single SNP present can encourage a mechanistic study to confirm our observations. Although this study presents strong evidence for the role of *MUC7* in CL/P, alternative and additional genes and regulatory elements could provide other possible mechanisms of clefting. This evidence promotes the concept that clefting and the development of CL/P have a strong genetic influence.

## Data Availability

The data that support the findings of this study are available in Sequence Read Archive at https://www.ncbi.nlm.nih.gov/bioproject/PRJNA700692, reference number PRJNA700692, and accession numbers SRX10054254, SRX10054253, SRX10054252, SRX10054250, SRX10054249.

